# Incorporating Public Values into Public Health Communications: Effects of Value Affirmations on Intentions to Vaccinate and Trust in the U.S.

**DOI:** 10.1101/2025.04.24.25326364

**Authors:** Christopher Wolsko, Elizabeth Marino

## Abstract

A set of COVID-19 pro-vaccine messages that aligned with key social values of vaccine-hesitant individuals was developed. Two separate experimental tests of these messages were conducted during the Spring of 2021 in the U.S., just as the public rollout of COVID-19 vaccines was getting fully underway. Unvaccinated adults in the U.S. (*n*=1,170) completed an online survey in which they were randomly assigned to read through one of several different pro-vaccine messages, followed by assessments of their future COVID-19 vaccine likelihood and level of trust in government, health care, and vaccines. Relative to a control condition, messages that affirmed individuals’ distrust in the government and concerns over restrictions on personal liberty resulted in *higher* vaccination likelihood and *greater* trust. The findings have implications for broadening the value frames of public health appeals and taking seriously how and why vaccine hesitancy acts as a powerful way for people to express their values and authentic social experiences.

## Introduction

Effective management of public responses to the COVID-19 pandemic was challenged by numerous factors, including the inherent uncertainty and severity of the evolving crisis, communication inefficiencies and inconsistencies (Paek & Hove, 2020; Sauer, Truelove, Gerste, & Limaye, 2021), propagation of mis- and disinformation (Ferreira Caceres, Sosa, Lawrence, Sestacovschi, Tidd-Johnson, Rasool et al., 2022) and low levels of trust between diverse members of the public, the biomedical and pharmaceutical industries, public health institutions, and government entities (Casselman-Hontalas, Adams-Santos, & Watkins-Hayes, 2024). Determining the most efficient and persuasive way to convey essential public health information is a perennial puzzle, and vaccine communication is no exception, as the pandemic brought into sharp relief. We are long past the assumption that people are essentially rational decision-makers when it comes to health care, or other domains (Kahneman & Tversky, 1979; Tversky & Kahneman, 1974, 1981), and research testing vaccination messaging techniques confirms this, demonstrating that simple fact-based communications are frequently ineffective (Hornsey, Harris, & Fielding, 2018; MacDonald, Butler, & Dube, 2018). Cognizant of this work, we set out during the initial public rollout of the COVID-19 vaccines in the U.S. in early 2021 to better understand some of the factors that would predict whether or not people decided to get vaccinated.

While research into public health messaging strategies has been dominated by a limited focus on gain vs. loss framing (Guenther, Gaertner, & Zeitz, 2020), a more recent, and promising, area of investigation has examined the effects of salient social identities and/or values present in messages (Amin, Bednarczyk, Ray, Melchiori, Graham, Huntsinger, et al., 2017; Chu et al., 2021; Limbu & Christopher McKinley, 2025). In their review of randomized controlled vaccine communication trials, Xia and Nan (2024) found that the most persuasive messages tended to be those that highlighted a shared identity between the message source and the audience, and a few studies have found that framing messages in terms of specific values that are similar to those of the audience may improve vaccine uptake (Heine & Wolters, 2021; Winters, Christie, Melchinger, Arias, Lirman, Thomson et al., 2024). This general strategy of pairing the social values in the message with the social values of the audience has also shown promise by reducing polarization and resistance to other contentious issues in the public square, including removing firearms as a suicide prevention strategy in the U.S. (Marino, Wolsko, Keys and Wilcox, 2018; Wolsko, Marino, & Keys, 2020) and taking conservation action in the face of climate change skepticism (Hurst & Stern, 2020; Wolsko, Ariceaga, & Seiden, 2016).

The mechanisms behind public health attitude shifts of this type may be usefully interpreted in light of persuasion research on matching effects. Research in this area indicates that attitude and behavior change will be greater when persuasive communications are congruent with the content, structure, and/or function of the recipient’s attitude in the relevant domain (Maio & Haddock, 2015; Watt, Maio, Haddock, & Johnson, 2009). That is, the use of specific identity or value frames to alter attitudes appears to be effective, at least in part, by virtue of its success in matching the self- or value-expressive functions of respondents’ attitudes.

In highly politicized and contentious domains, such as with COVID-19 policy and vaccine attitudes in the U.S., individuals’ expressions of their attitudes are about far more than a particular attitude object – a vaccine in this case. Instead, attitudes serve important self-expressive functions, acting as vehicles for conveying social identity, lived experiences, and important social or moral values (Kahan, 2013). From this perspective, a person’s expression of an attitude against getting a COVID-19 vaccine may be seen as an affirmation of a particular social experience or value – for example, distrust of the government or of the health care system.

Just as an attitude may persist because it meets a functional need, so to, “attitude change occurs to meet a functional need” (Watt et al., 2009, p. 194). In the present context, we expected that the attitudes of the unvaccinated may change in the pro-vaccination direction under conditions in which specific self- or value-expressive functions are actually being fulfilled by agreeing with a pro-vaccine agenda (rather than being stifled, threatened, or deemed irrational by agreeing with an anti-vaccine agenda).

To determine the content of these value-expressive functions, we conducted a series of focus group interviews with vaccine-hesitant adults in the U.S. during the late winter and early spring of 2021. We sought to understand their most pressing concerns and to probe the perceptions of risk, cultural identities, and moral values that underpinned their COVID-19-related attitudes and behaviors. From these interviews, and from our observations of the contentious public discourse around vaccines at the time, we developed a set of COVID-19 pro-vaccine messages that aligned with key social values that appeared to be in play for peoples’ decision to vaccinate, especially for those who were hesitant. We then conducted two separate experimental tests of the effects of these messages on the likelihood of non-vaccinated individuals getting a COVID-19 vaccine in the future, and on their level of distrust in government, health care, and vaccines. We present these experimental findings below.

## Materials and Methods

### Participants and Procedural Overview

Two separate surveys were conducted during the Spring of 2021, just as the first major waves of people in the U.S. were getting vaccinated. All participants were sampled from the Amazon MTurk system and completed one of two short 10-15 minute surveys in exchange for $1.00. MTurk is an online labor market that has been widely utilized by survey researchers in psychology and other social sciences, and samples obtained are generally demographically diverse, representative of the U.S. population, and display strong psychometric properties (e.g. test-retest reliability, experimental replication) (Buhrmester, Kwang, & Gosling, 2011; Berinsky, Huber, & Lenz, 2012). All individuals were instructed to complete the survey only if they had *not* received a vaccination for COVID and they had *not* made an appointment to do so.

Survey 1 was conducted on April 1, 2021, and had 371 participants. The sample was relatively balanced in gender (48.8% male, 51.2% female); majority Caucasian (66.8% White, 15.1% Black/African American, 5.1% Latino/Hispanic, 10.5% Asian American, 0.8% Pacific Islander; 0.8% American Indian / Alaska Native); diverse in age (*M* = 38.29, *SD* = 11.58; range: 18 to 76 years old); diverse in highest educational attainment (0.5%, some high school; 6.5%, high school diploma or GED; 23.2%, some college or associates degree; 45.3%, bachelor’s degree; 24.5%, master’s degree or higher); and representative of the U.S. population in household income (9.7%, less than $25,000; 24.5% from $25,000 to $49,999; 22.9% from $50,000 to $74,999; 23.2% from $75,000 to $99,999; 19.7%, $100,000 or more).

Survey 2 was completed on May 24, 2021, and had 799 participants. Demographics were similar to Survey 1, except for a slightly greater percentage of male respondents (58.5% male, 41.5% female). The majority were Caucasian (66.7% White, 20.2% Black/African American, 4.9% Latino/Hispanic, 5.6% Asian American, 0.4% Pacific Islander; 1.6% American Indian / Alaska Native); diverse in age (*M* = 36.95, *SD* = 10.86; range: 18 to 80 years old); diverse in highest educational attainment (0.4%, some high school; 6.9%, high school diploma or GED; 20.8%, some college or associates degree; 49.9%, bachelor’s degree; 22.1%, master’s degree or higher); and representative of the U.S. population in household income (9.9%, less than $25,000; 24.9% from $25,000 to $49,999; 31.2% from $50,000 to $74,999; 19.3% from $75,000 to $99,999; 14.6%, $100,000 or more).

Participants in both surveys were first randomly assigned to read information either from a control condition or from one of several vaccine messaging conditions. Following, individuals in both surveys responded to the following assessments in this order: COVID vaccine intentions; distrust in vaccines, government, and the health care system; and demographics. Condition and key variable names are italicized in the descriptions that follow.

#### Ethics Statement

These anonymous survey responses were collected as part of a program evaluation of COVID-19-related public health communications at the Deschutes County Health Department in 2021. The data was archived after this program evaluation and then later approved for analysis as research in 2022 by the Human Research Protection Program and Institutional Review Board at Oregon State University (IRB-2022-1357).

### Vaccine Messaging Conditions

The dominant themes from our discourse analyses were distrust in sociopolitical and medical institutions, concerns about one’s ability to exercise personal liberty and freedom of choice (including whether or not to get a vaccine), and desires to protect and reconnect with family. From this analysis, we developed four distinct slogans: (1) “You don’t have to trust the government, here are the facts” was designed to affirm individuals’ social experience of feeling like politicians and governmental organizations are not trustworthy; (2) “Everybody wants to get back to their families” was designed to affirm the importance of family as an identity and of people’s concerns about maintaining normal family life; and (3) “It’s your life, it’s your choice” and (4) “COVID and politicians: Don’t tread on me” were designed to affirm the values of personal liberty and freedom.

In Survey 1, participants were randomly assigned to read through one of 4 different messages: *control*; *families*; *government*; and *choice* (see Appendix for the full text of each message). In the *control* condition, participants read the following statement: “First, consider the following information about the COVID-19 vaccines from physicians at the Mayo Clinic and the Centers for Disease Control and Prevention (CDC).” They were then presented with one page of actual information from these organizations that detailed the safety, efficacy, and rationale for the COVID-19 vaccines (see Appendix). In the other 3 conditions, just prior to reading this same information about the COVID-19 vaccines from the Mayo Clinic and the CDC, participants were exposed to one of the slogans, described above, in large, bold font. In the *families* condition, the slogan read, “Everybody wants to get back to their families.” In the *government* condition, the slogan said, “You don’t have to trust the government, here are the facts.” In the *choice* condition, the slogan read, “It’s your life, it’s your choice.” Participants in Survey 2 were randomly assigned to one of these same four conditions *or* to a fifth, the *liberty* condition, which read, “COVID and politicians: Don’t tread on me,” before being shown the same information from the Mayo Clinic and the CDC.

### COVID Vaccine Attitudes

Participants in both surveys responded to two items that assessed the likelihood of getting a COVID vaccine in the future. The first item asked participants, “How likely are you to get an FDA authorized vaccine for COVID-19, once it is available to you for free?” Participants responded to this item on a 6-point scale, ranging from 1, *I will definitely NOT get the vaccine,* to 6, *I am certain that I’ll get the vaccine*. Participants were assigned a *vaccine likelihood* score, based on their response to this item.

Additionally, in Survey 2, participants with children under the age of 18 were asked, “How likely is it that you will have your child get an FDA authorized vaccine for COVID-19, once it is available for free? If you have more than one child under 18, answer for your youngest.” Participants responded to this item on a 6-point scale, ranging from 1, *I will definitely NOT have my child get the vaccine*, to 6, *I am certain that I’ll have my child get the vaccine.* Participants in Survey 2 were assigned a *child vaccine likelihood* score, based on their response to this item.

The second type of vaccine likelihood item was similar to an assessment that was widely utilized at the time, including by the Kaiser Family Foundation, an American non-profit that is one of more prolific assessors of public health attitudes. Participants were asked, “When an FDA authorized vaccine for COVID-19 is available to you for free, do you think you will…?” and instructed to select one of four response options: probably get vaccinated as soon as possible; wait and see; get vaccinated only if it’s required for my work or travel; or definitely not get vaccinated. Each participant received a score on this categorical variable, *vaccine intention*.

### Distrust

Next, participants responded to three items designed to assess their level of distrust in vaccines (“I don’t trust vaccines in general”), the government (“I don’t trust the government to make sure the COVID vaccines are safe and effective”), and the health care system (“I don’t trust the health care system”). Responses were provided on a 7-point scale, ranging from 1, *strongly disagree*, to 7, *strongly agree*. The items were strongly intercorrelated, and a principal components factor analysis suggested a clear one-factor solution, so we decided to combine them into a single index of *distrust* (α = .87 for Survey 1; α = .83 for Survey 2), by taking the mean for each participant across all three items, with higher numbers indicating reflecting *less* trust in vaccines, the government, and health care.

### Demographics

Finally, participants responded to a series of demographic questions, including gender, age, education, annual income, race/ethnicity, and political orientation. Descriptions of these variables were presented above, with the exception of political orientation, which was assessed by asking participants to indicate the degree to which they identified as more liberal or more conservative. Responses were obtained on a 7-point scale (1 = *extremely liberal*; 2 = *moderately liberal*, 3 = *slightly liberal*; 4 = *moderate*; 5 = *slightly conservative*; 6 = *moderately conservative*; 7 = *extremely conservative*), and each individual received a *political orientation* score based on their response to this item.

## Results

### Demographic Correlates of Vaccine Attitudes

We first examined the extent to which, across all conditions of the surveys, our primary dependent variables of *vaccine likelihood* and *vaccine intention* were associated with participants’ demographic characteristics. Results indicated that political orientation, education, and income were the only demographics to emerge systematically as reliable predictors of vaccine attitudes. In the service of this brief report, we highlight just a few of the findings. Participants who expressed a greater likelihood of obtaining a COVID-19 vaccine (higher scores on *vaccine likelihood*) also reported having a higher income (Survey 1, *r*(369)=.16, *p*=.001; Survey 2, *r*(792)=.11, *p*=.002), being more educated (Survey 1, *r*(369)=.10, *p*=.049; Survey 2, *r*(792)=.13, *p*<.001), and being more politically liberal (Survey 1, *r*(368)=-.31, *p*<.001; Survey 2, *r*(790)=-.23, *p*<.001). To gain a greater focus on the effects of our messaging over and above significant demographic predictors, our models in the next section include income, education, and political orientation as covariates.

### Experimental Effects of Messages on Vaccine Attitudes

The means (and frequencies, where relevant) for the vaccine attitude variables as a function of condition and survey are presented in Table 1. Our predictions were that messages that affirmed individuals’ social experience and values would precipitate greater probability of vaccination. Thus, we wanted to compare each messaging condition to the control condition, resulting in a set of three condition contrasts for Survey 1 (*control vs. government*, *control vs. family*, and *control vs. choice*) and four contrasts for Survey 2 (the same contrasts for Survey 1 and an additional for *control vs. liberty*). We conducted a series of regression analyses in which each of the 3 dependent variables assessing vaccination attitudes (*vaccine likelihood*, *child vaccine likelihood, and vaccine intention*), was regressed, in turn, on one of the condition contrasts (Model 1: *control vs. government*; Model 2: *control vs. family*; Model 3: *control vs. choice*, and Model 4: *control vs. liberty*), along with political orientation, income, and education. In reporting our findings, we focus solely on the significance of coefficients for the condition contrast terms.

**Table 1.** Means and Frequencies for Vaccination Attitudes by Condition and Survey Survey 1 Vaccine Likelihood.

| *Condition* | *Vaccine Likelihood* | *Vaccine Intention* |  |  |  | *Distrust* |
| --- | --- | --- | --- | --- | --- | --- |
|  |  | Get it ASAP | Wait and See | Only if Required | Definitely Not |  |
| Control (n=93) | 3.82 | 49.5% | 21.5% | 10.8% | 18.3% | 3.91 |
| Government (n=93) | 4.33 | 65.6% | 19.8% | 7.3% | 7.3% | 3.65 |
| Families (n=96) | 4.33 | 58.1% | 24.7% | 11.8% | 5.4% | 3.51 |
| Choice (n=89) | 4.15 | 55.1% | 25.8% | 7.9% | 11.2% | 3.92 |

Survey 2
| *Condition* | *Vaccine Likelihood* | *Child Vaccine Likelihood* | *Vaccine Intention* |  |  |  | *Distrust* |
| --- | --- | --- | --- | --- | --- | --- | --- |
|  |  |  | Get it ASAP | Wait and See | Only if Required | Definitely Not |  |
| Control (n=162) | 3.31 | 3.04 | 40.7% | 29.6% | 17.9% | 11.7% | 4.37 |
| Government (n=155) | 3.78 | 3.32 | 45.2% | 27.1% | 18.7% | 9.0% | 4.26 |
| Families<br>(*n*=170) | 3.46 | 3.23 | 39.4% | 30.0% | 15.9% | 14.7% | 4.23 |
| Choice<br>(*n*=156) | 3.51 | 3.25 | 39.0% | 24.0% | 20.5% | 16.4% | 4.30 |
| Liberty<br>(*n*=163) | 3.71 | 3.55 | 48.5% | 34.4% | 8.0% | 9.2% | 3.84 |

*Notes:* Means are presented for *vaccine likelihood*, *child vaccine likelihood*, and *distrust*. Likelihood variables were assessed on a 6-point scale, with higher numbers indicating greater likelihood. *Distrust* was assessed on a 7-point scale, with higher numbers indicating more distrust of health care, government, and vaccines. Percentages represent relative frequencies within condition for *vaccine intention*.

In Survey 1, the *control vs. government* coefficient was significant in Model 1, *B*=.33, *SE*=.13, *t*(184)=2.58, *p*=.011; as was the *control vs. family* coefficient in Model 2, *B*=.28, *SE*=.13, *t*(187)=2.21, *p*=.028. These findings indicated that participants reported being more likely to get a COVID vaccine in the *government* condition and in the *family* condition than in the *control*. The *control vs. choice* coefficient in Model 3 did not approach significance, *B*=.12, *SE*=.13, *t*(179)=0.93, *p*=.352.

In Survey 2, the *control vs. government* coefficient was significant in Model 1, *B*=.21, *SE*=.09, *t*(313)=2.38, *p*=.018; as was the *control vs. liberty* coefficient in Model 4, *B*=.17, *SE*=.09, *t*(322)=2.00, p=.047. These findings indicated that participants reported being more likely to get a COVID vaccine in the *government* condition and in the *liberty* condition than in the *control*. The *control vs. family* coefficient in Model 2 was not significant, *B*=.07, *SE*=.09, *t*(329)=0.80, *p*=.422; nor was the *control vs. choice* coefficient in Model 3, *B*=.09, *SE*=.09, *t*(306)=0.98, *p*=.326.

#### Child Vaccine Likelihood

In Survey 2, only the *liberty* coefficient in Model 4 was a significant predictor of *child vaccine likelihood*, *B*=.23, *SE*=.10, *t*(296)=2.40, *p*=.017, indicating that parents reported being more likely to have their child get a COVID vaccine in the *liberty* condition than in the *control*. The *control vs. government* coefficient in Model 1 was not significant, *B*=.13, *SE*=.10, *t*(289)=1.36, *p*=.176; nor was the *control vs. family* coefficient in Model 2, *B*=.09, *SE*=.10, *t*(306)=0.98, *p*=.328; nor was the *control vs. choice* coefficient in Model 3, *B*=.09, *SE*=.10, *t*(306)=0.91, *p*=.363.

#### Vaccine Intention

As our *vaccine intention* variable is technically nominal with four categories, we decided to simplify analyses by recategorizing responses into two levels: 0, if participants indicated that they were definitely not going to get a shot, or get one only if they were required to do so; vs. 1, if participants indicated that they were probably going to get vaccinated as soon as possible, or that they would wait and see. Binomial logistic regressions were then conducted with this new dichotomous dependent variable as the outcome, and the same sets of predictors as described in the previous regression analyses.

In Survey 1, the *control vs. government* coefficient was a significant predictor of *vaccine intention* in Model 1, *B*=.48, *SE*=.19, *Wald*=6.12, *p*=.013, *Exp(B)*=1.61; as was the *control vs. family* coefficient in Model 2, *B*=.40, *SE*=.19, *Wald*=4.50, *p*=.034, *Exp(B)*=1.48. These findings indicated that participants reported being more likely to get a COVID vaccine or to wait and see in the *government* condition and in the *family* condition than in the *control*. The *control vs. choice* coefficient in Model 3 was not significant, *B*=.24, *SE*=.19, *Wald*=1.63, *p*=.202, *Exp(B)*=1.27.

In Survey 2, only the *liberty* coefficient in Model 4 was a significant predictor of *vaccine intention*, *B*=.35, *SE*=.14, *Wald*=6.43, *p*=.011, *Exp(B)*=0.71, indicating that participants reported being more likely to get a COVID vaccine or wait and see in the *government* condition than in the *control*. The *control vs. government* coefficient in Model 1 was not significant, *B*=.04, *SE*=.13, *Wald*=0.10, *p*=.754, *Exp(B)*=0.96; nor was the *control vs. family* coefficient in Model 2, *B*=.03, *SE*=.12, *Wald*=0.06, *p*=.810, *Exp(B)*=0.97; nor was the *control vs. choice* coefficient in Model 3, *B*=.18, *SE*=.12, *Wald*=2.10, *p*=.147, *Exp(B)*=0.84.

### Experimental Effects of Messages on Distrust

The means for the distrust variable as a function of condition and survey are presented in Table 1. Message condition effects on the index of *distrust* were examined using the same sets of regression models as for vaccine likelihood. In Survey 1, the *control vs. government* coefficient was a marginally significant predictor of *distrust* in Model 1, *B*=.25, *SE*=.15, *t*(187)=1.69, *p*=.093; and the *control vs. family* coefficient was significant in Model 2, *B*=.30, *SE*=.15, *t*(184)=1.99, *p*=.048. These findings indicated that participants reported *lower* levels of distrust in the *family* condition than in the *control*, and marginally lower distrust in the *government* condition than in the *control*. The *control vs. choice* coefficient in Model 3 was not significant, *B*=.04, *SE*=.16, *t*(179)=0.27, *p*=.790.

In Survey 2, only the *liberty* coefficient in Model 4 was a significant predictor of *distrust*, *B*=-.22, *SE*=.09, *t*(322)=-2.50, *p*=.013, indicating that participants reported lower levels of distrust in the *liberty* condition than in the *control*. The *control vs. government* coefficient in Model 1 was not significant, *B*=-.02, *SE*=.09, *t*(313)=-0.25; *p*=.801, nor was the *control vs. family* coefficient in Model 2, *B*=.06, *SE*=.09, *t*(329)=0.69, *p*=.504; nor was the *control vs. choice* coefficient in Model 3, *B*=.02, *SE*=.09, *t*(306)=0.23, *p*=.820.

## Discussion

These findings provide some support for our hypothesis that public health messaging that affirms peoples’ social experiences and values would be more influential than standard information-based messaging alone. Across outcome variables, the most impactful messages were those that affirmed distrust in the government, the importance of family, and value of personal liberty. Here, we briefly summarize key results and discuss implications for future research in this area.

In Survey 1, relative to the control condition, validating peoples’ wariness about the government increased individuals’ expressed likelihood of getting vaccinated, and, ironically, also marginally *increased* trust in the government, health care, and vaccines. Emphasizing the importance of family in peoples’ lives also significantly increased the likelihood of getting vaccinated and trust in the government, health care, and vaccines.

Vaccine attitudes at two months later, in Survey 2, were less influenced by message content. Emphasizing governmental distrust continued to shift individuals’ behavioral intentions in the direction of greater vaccine likelihood, but highlighting the importance of family was not a significant predictor for any of our variables. In contrast, a new condition that emphasized liberty concerns resulted in higher vaccine likelihood, both for oneself and for one’s children, as well as greater trust in the government, health care, and vaccines in general.

Consistent with prior work on attitude change, the present findings support the notion that health care attitudes and behaviors are much more than the outcome of rational deliberation – they function as profound expressions of individuals’ personal and cultural values (Amin et al., 2017; Chu et al., 2021; Limbu & Christopher McKinley, 2025). While direct communication of public health information to the public remains a laudable and essential goal, how individuals or subcultural constituencies interpret this information and how they choose to act or not act on the basis of it will always remain socially-motivated— a function of what is deemed important, moral, and identity-affirming. Because such motivations are so profoundly influential in human culture and psychology, we argue that it is immensely important to promote inclusivity and tolerance of multiple moral and ideological frameworks, and to appeal to the functions of attitudes from multiple perspectives.

Although the framing effects investigated here may have substantial effects on behaviors such as vaccination, we caution against using evidence such as this as a simple tool of persuasion. In our work studying COVID-19 responses, our goals were to listen and categorize people’s important values and perceptions of the risk as a means to enrich the multiplicity of public discourse, and to take seriously how and why vaccine hesitancy acts as a reasonable way to express the situatedness of one’s personal and collective social life. Understanding and empathizing with the experiences of a vaccine-hesitant public is likely to make communication around health policy issues less defensive, less volatile, and more open to new information.

## Data Availability

Data can be accessed in the Supporting Information for this research article.

## Acknowledgements

This research was generously supported by grants from the Deschutes County Health Department, and the Roundhouse Foundation.

## Notes

### Competing Interest Statement

The authors have declared no competing interest.

### Funding Statement

This research was supported by grants from the Deschutes County Health Department and the Roundhouse Foundation. Funds were provided to the authors to conduct a program evaluation to better understand how to communicate with the public about COVID-19 vaccines and other socioculturally contentious issues in the U.S.

### Author Declarations

Human Research Protection Program and Institutional Review Board, Oregon State University.

